# Implementing blended cognitive behavioral therapy for severe fatigue in routine hospital care for diffuse glioma patients: a qualitative study

**DOI:** 10.64898/2026.10.30.26366910

**Authors:** Ilias Echaoui, Maxine Gorter, Marieke E.C. Blom, Johanna M. Niers, Philip C. De Witt Hamer, Hans Knoop, Linda Douw, Martin Klein, Marian Smeulers

## Abstract

**Background:** Severe fatigue is common in diffuse glioma patients and can persist after tumor treatment. Fatigue-tailored blended cognitive behavioral therapy (bCBT) has been shown to reduce severe fatigue in glioma patients. This study aimed to identify barriers and facilitators to implementing bCBT for severe fatigue in glioma care and to develop implementation strategies tailored to the neuro-oncology context.

**Methods:** A qualitative study was conducted among healthcare professionals involved in glioma care, including neurologists, nurse practitioners, and (neuro)psychologists. Semi-structured interviews were conducted with a topic list guided by the Consolidated Framework for Implementation Research. Data were analyzed using a combined deductive and inductive approach. Implementation strategies were developed by integrating identified barriers and facilitators, participant suggestions, and research team expertise.

**Results:** Twenty-six healthcare professionals from several Dutch neuro-oncological centers participated in this study. Key themes of implementation included the perceived need for fatigue treatment with bCBT, referral to psychological care, knowledge on patient eligibility for bCBT, awareness of fatigue and bCBT, confidence in its evidence, capacity constraints, digital platform access and financial responsibility. Implementation strategies were tailored to identified barriers across three phases: identifying fatigue, referring diffuse glioma patients and treating severe fatigue with bCBT.

**Conclusions:** Implementation of bCBT for severe fatigue in glioma care requires more than evidence of effectiveness. Successful integration into routine hospital care will depend on timely fatigue identification, clear referral responsibilities, appropriate patient selection, trained professionals and alignment with local organizational and financial conditions. Center-specific implementation strategies may facilitate integration of bCBT into neuro-oncology care.

**Key points:**

- Awareness of fatigue and bCBT together with eligibility guidelines for bCBT are key for adoption.
- Referral pathways, financial responsibility and limited capacity may hinder implementation.
- Integration into routine hospital care will likely require center-specific strategies.

**Importance of study:** Blended cognitive behavioral therapy (bCBT) is effective in reducing severe fatigue in patients with diffuse glioma. This qualitative study was conducted as a pre-implementation step from the perspectives of neurologists, nurse practitioners, and (neuro)psychologists to support adoption of bCBT in neuro-oncology care. Barriers and facilitators identified were related to the need for fatigue reduction, awareness of fatigue and bCBT, confidence in its evidence, patient suitability, referral pathways, psychological care capacity, and financial responsibility. Implementation needs differed across centers according to existing fatigue assessment practices, care pathways, organizational structures, and available digital platforms. These findings indicate that implementation is unlikely to be achieved by a single uniform strategy. Instead, center-specific approaches aligned with existing workflows are needed.

**Lay summary:** Fatigue is the most common and burdensome symptom for people with brain tumors (gliomas). A cognitive-behavioral treatment combining online and in-person therapy, has shown to help, but hospitals have not widely adopted it. We interviewed neurologists, nurse practitioners and psychologists to understand what helps or hinders its use in hospital care, then developed strategies to address the challenges identified. Healthcare professionals recognized a clear need for fatigue reduction, but uptake in routine care may be hindered by limited awareness of fatigue and the treatment, staffing and resource constraints and unclear financial responsibility. Different hospitals will likely need tailored approaches for successful adoption.

## Introduction

Fatigue is among the most prevalent and debilitating symptoms reported by patients with diffuse glioma.^1,2^ Up to 96% of patients experience severe fatigue at some point during the disease trajectory.^3–5^ Acknowledging and treating severe fatigue could significantly improve health-related quality of life (HRQoL) in glioma patients.^2,6–8^ Although fatigue constitutes a major symptom burden, no pharmacological treatments have been proven to be effective.^9–12^ In contrast, blended cognitive behavioral therapy (bCBT) specifically aimed at fatigue has recently demonstrated effectiveness in reducing severe fatigue in glioma patients.^13^ However, it has not yet been integrated into routine hospital care for this patient population. This implementation gap is clinically important, as it may prevent patients with severe fatigue from receiving evidence-based care.

Fatigue often emerges during the disease course in patients with diffuse glioma and frequently persists after completion of tumor treatment.^2–5^ This suggests that disease- and treatment-related factors may initiate fatigue, while post-treatment behavioral, psychological, and physiological factors may contribute to its persistence.^14,15^ The distinction is clinically relevant because persistent severe fatigue is consistently associated with symptoms of depression, anxiety, sleep disturbance, and substantial reductions in HRQoL.^2,3,8,16^ Despite this, fatigue in patients with diffuse glioma is often interpreted as an inevitable component of the disease course, a consequence of oncological treatment, or part of the cognitive impairment commonly observed in this population. As a result, it may not always be recognized as a distinct symptom requiring targeted intervention.^6,7,17^ Furthermore, diffuse glioma represents a patient population with a relatively limited life expectancy following treatment, emphasizing the importance of timely identification and management of their fatigue symptoms.^18,19^

Cognitive behavioral therapy (CBT) is an established approach for targeting severe fatigue and has demonstrated effectiveness across various cancer and non-oncological populations with cognitive impairment, including multiple sclerosis, stroke, and acquired brain injury.^20–24^ Fatigue-tailored CBT interventions specifically target perpetuating factors such as sleep disturbance and maladaptive cognitions, including catastrophizing thoughts, and have been shown to significantly reduce severe fatigue across populations.^14^ bCBT combines CBT with online modules completed at home with therapist feedback. Importantly, a randomized controlled trial recently showed that bCBT addressing fatigue indeed reduces fatigue and improves HRQoL in patients with diffuse glioma.^13^ Moreover, this blended format has the potential to improve accessibility and flexibility and reduce healthcare costs while maintaining the continuity of care and improving the HRQoL of diffuse glioma patients.^13,25^

Despite its effectiveness, bCBT has not yet been implemented in routine hospital care for patients with diffuse glioma, potentially limiting access to evidence-based care for patients with severe fatigue. Implementation research provides a way to explore contextual factors across different tertiary centers while also raising awareness of bCBT to accelerate the uptake of the intervention in standard hospital care.^26^ The aim of this study was to identify expected barriers and facilitators to the implementation of fatigue-tailored bCBT from the perspective of healthcare professionals from tertiary centers treating diffuse glioma patients, and to translate these findings, using ERIC matching strategies, into tailored implementation recommendations for routine clinical practice.

## Methods

### Study design

This qualitative study was conducted by Amsterdam University Medical Center (Amsterdam UMC) and consisted of a pilot and two sequential phases. The pilot phase served to assess and refine the topic list and interview procedure. The aim of the first phase was to identify barriers and facilitators to the implementation of bCBT for fatigue in routine hospital care for patients with diffuse glioma. The second phase focused on the development of targeted implementation strategies. The study was conducted and reported in accordance with the Consolidated Criteria for Reporting Qualitative Research (COREQ) (Supplementary data).^27^

### Study population

The study population consisted of healthcare professionals from 8 Dutch neuro-oncology centers (Table 1). The study aimed to include at least one nurse practitioner, one neurologist, and one psychologist per center to capture perspectives from all healthcare professionals that would be involved in identifying, referring, and treating glioma patients with bCBT. Purposeful sampling was used to include both professionals who were supportive of the implementation of bCBT and those who were more critical or hesitant.^28^ During each interview, participants were asked to suggest a colleague of the other predetermined disciplines. This approach was employed to ensure a balanced range of views while minimizing selection bias. This process continued until saturation was reached. Participants were recruited via email. All participants received information about the study procedures. Informed consent was obtained from every participant by email and this study was approved by the medical ethical committee of Amsterdam UMC (METc 2026.0223).

**Table 1.**
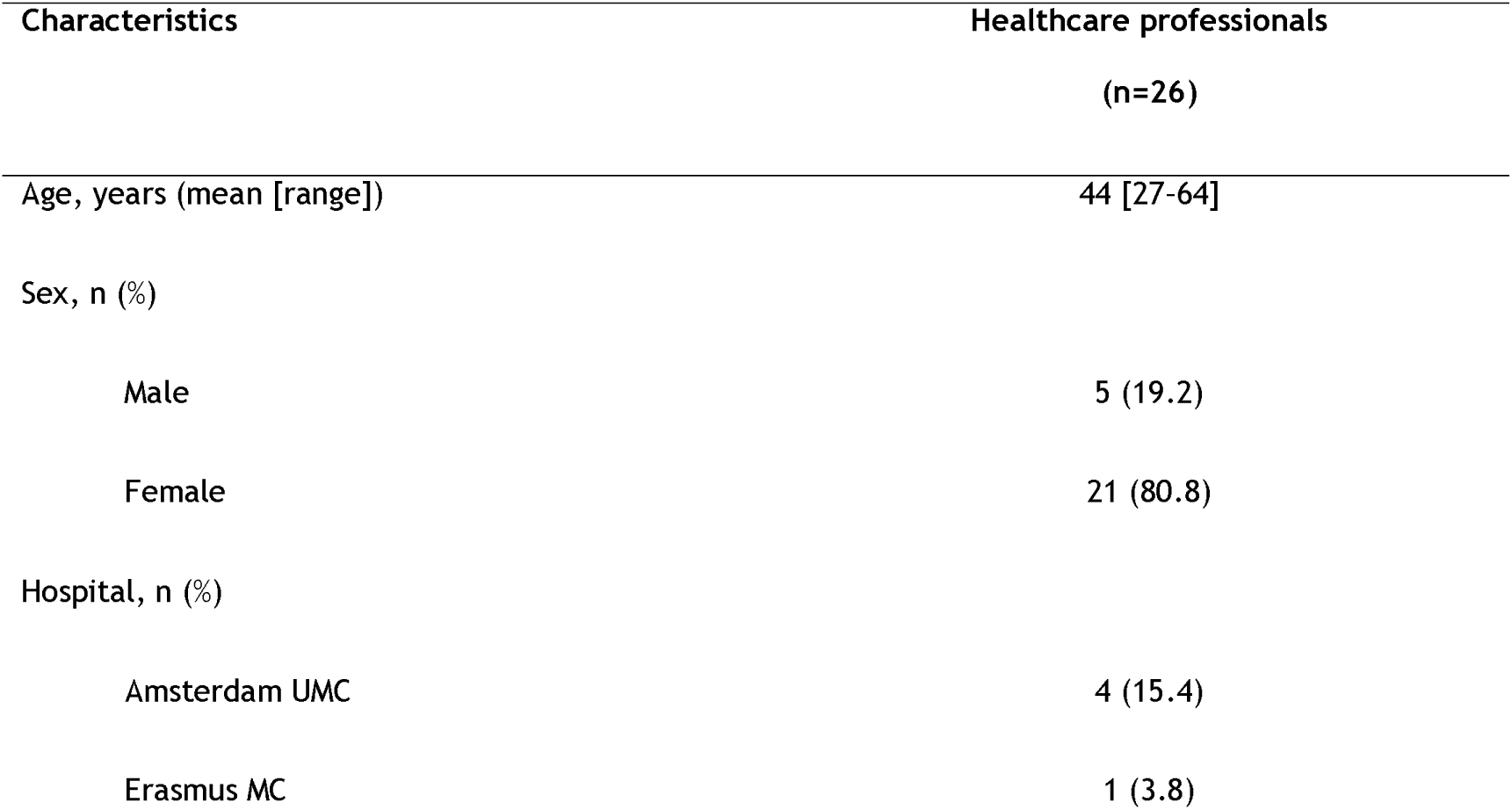
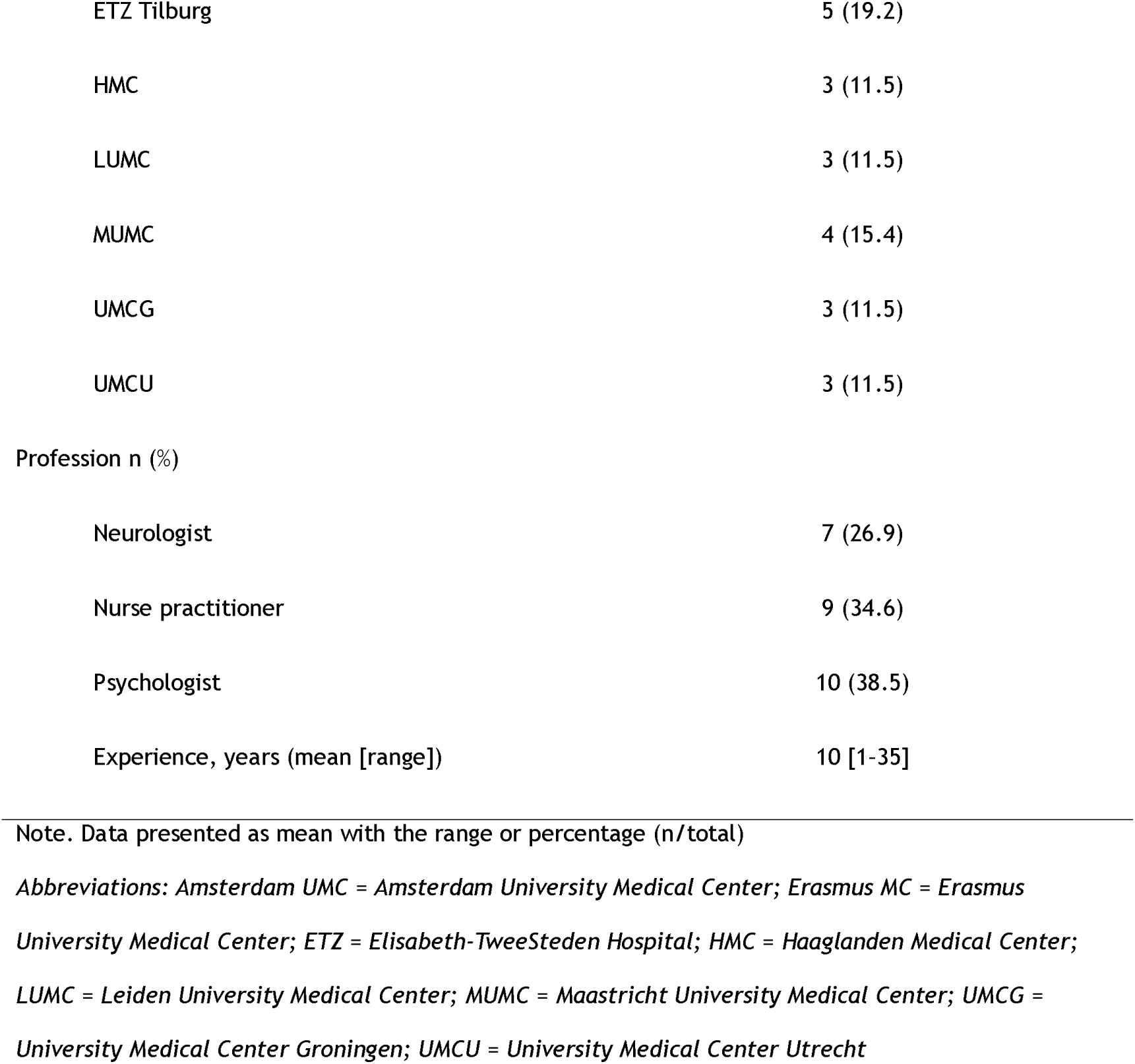
Characteristics of the healthcare professionals.

**Table 2.** Barriers and facilitators to the implementation of bCBT, organized by theme and corresponding CFIR constructs.

| Theme | CFIR constructs | Barriers | Facilitators |
|---|---|---|---|
| Need for fatigue reduction with bCBT | Relative Advantage; Available Resources; Patient Needs & Resources | • Unclear added value over existing CBT | • Recognized unmet need<br>• Limited alternative options |
| Awareness as a prerequisite for adoption | Knowledge & Beliefs about the Intervention; Access to Knowledge & Information; Structural Characteristics; Patient Needs & Resources | • Limited familiarity with bCBT and its fatigue-tailored content<br>• Underlying mechanisms of fatigue considered difficult to understand<br>• Fatigue not routinely assessed<br>• Patients do not always mention fatigue<br>• Limited consultation time | • Multifactorial nature of fatigue widely acknowledged<br>• Psychologists routinely assess fatigue<br>• Nurse practitioners address fatigue |
| Evidence as a lever for change | Evidence Strength & Quality; Knowledge & Beliefs about the Intervention | • A single RCT may be insufficient to change guidelines<br>• Anticipated skepticism among colleagues | • Positive attitude due to RCT effectiveness |
| Matching the intervention to the patient | Relative Advantage; Complexity; Patient Needs & Resources | • Uncertainty about positioning bCBT relative to other options<br>• Suitability for WHO high-grade glioma is complex | • Agreement that patients with severe, persistent, debilitating fatigue are suitable |
| Pathways to psychological care | Structural Characteristics; Implementation Climate; Compatibility | • Limited in-hospital referral option<br>• Fatigue rarely the primary referral reason | • Close collaboration with psychological care<br>• Pre-existing link with neuropsychologists assessing glioma patients |
| Capacity constraints | Available Resources; Structural Characteristics; Implementation Climate | • Psychology departments at full capacity<br>• Waiting lists of several months | • Limited number of eligible patients<br>• Urgent cases prioritized<br>• Waiting times of weeks |
| Funding the foundation | Costs; Available Resources; Compatibility | • Implementation difficult without clear cost arrangements<br>• Cost-efficiency questioned for a small subgroup<br>• App subscription used (Therapieland vs. MindDistrict) | • Costs not perceived as a problem<br>• Training costs and time considered manageable |

*Note*. *Abbreviations: CFIR = Consolidated Framework for Implementation Research; bCBT = blended cognitive behavioral therapy; CBT = cognitive behavioral therapy; RCT = randomized controlled trial.*

### Blended Cognitive Behavioral Therapy

This study explored the potential implementation of bCBT. bCBT is a 12-week intervention consisting of five therapist-led sessions, delivered either face-to-face or via secure video consultation, and five to eight structured online therapy modules.^29^ The intervention targets fatigue-perpetuating factors and is delivered by trained cognitive behavioral therapists with expertise in fatigue-tailored interventions. The intervention is supported by an interactive online platform which enables patient engagement and therapist feedback. A detailed description of the intervention and treatment modules has been published previously.^29^

### Data collection

Semi-structured interviews were conducted via video call using Microsoft Teams. A topic list (see Supplementary Figure 1) was used to ensure consistency across interviews while allowing flexibility for in-depth exploration of participants’ perspectives, and was developed using the Consolidated Framework for Implementation Research (CFIR).^30^ CFIR is a comprehensive, theory-informed framework that integrates evidence-based implementation theories and provides a structured approach to identifying determinants of implementation. The framework comprises five major domains: 1) intervention characteristics, 2) inner setting, 3) outer setting, 4) characteristics of individuals, and 5) the implementation process. Within each domain, CFIR specifies multiple constructs representing contextual factors relevant to implementation.^30^ Although CFIR 2.0 is the updated version, CFIR 1.0 was used because the implementation strategies were based on the original CFIR 1.0 constructs, ensuring methodological consistency. Furthermore, it provided a clear and suitable framework for examining implementation factors from the perspectives of healthcare professionals and the healthcare organization. These CFIR constructs were operationalized by translating them into interview topics, enabling a comprehensive assessment of potential barriers and facilitators to the implementation of bCBT. Additions and revisions were made after the pilot interviews to enhance the relevance and quality of the topic list.

Semi-structured interviews were conducted between March and September 2026 by IE and MG. The interviews lasted approximately 30 minutes. All interviews were audio-recorded and transcribed by IE.

### Data analysis

#### Barriers and facilitators

Data analysis was performed through thematic analysis in parallel with the interviewing process by IE and MG. For thematic analysis, MAXQDA 24 was used.^31^ Thematic analysis is a method of identifying, analyzing and reporting patterns and consists of five steps: compiling, disassembling, reassembling, interpreting and concluding.^32^ Coding was done deductively using the predefined CFIR constructs and inductively by allowing codes to emerge from the data. MG independently coded 20% of the interviews during the data collection period for coding consistency throughout the study. Any discrepancies were discussed and resolved to 100% agreement. Themes were discussed and refined with a group of experts in neuro-oncology, neuropsychology, and implementation science.

#### Implementation strategies

Implementation strategies were developed using the ZonMw implementation framework, which links CFIR-based barriers to implementation strategies from the Expert Recommendations for Implementing Change (ERIC).^33,34^ The ZonMw framework provided targeted implementation strategies across several categories including informational, educational, organizational, and facilitative strategies.^34^ In addition, implementation suggestions provided by participants were taken into account, as these offered practical solutions to center-specific challenges that may also be encountered in hospitals with similar organizational structures. All implementation strategies and recommendations were discussed with the same expert group to assess their clinical relevance and feasibility. The strategies are presented according to three phases representing the clinical pathway for glioma patients: Phase 1: Identifying fatigue, Phase 2: Referring diffuse glioma patients, and Phase 3: Treating severe fatigue with bCBT.

## Results

A total of 26 participants were interviewed, including 7 neurologists, 10 psychologists and 9 nurse practitioners. Most participants were recruited from Elisabeth-TweeSteden Hospital, which contributed 5 participants (19.2%) as shown in Table 1. The majority of participants were female (80.8%). The mean age was 44 years, and participants had an average of 10 years of experience in their current profession.

### Barriers and facilitators

Seven themes were identified: need for fatigue reduction with bCBT, awareness as a prerequisite for adoption, evidence as a lever for change, matching the intervention to the patient, pathways to psychological care, capacity constraints, and funding the foundation. Themes encompassed multiple CFIR constructs, which are presented in brackets next to each theme. Each theme is described and supported by quotes from the participants.

#### Need for fatigue reduction with bCBT (relative advantage, available resources, patient needs and resources)

Many healthcare professionals, particularly nurse practitioners and neurologists, but also some psychologists, indicated that a small but clinically relevant subgroup of patients experience an unmet need for their severe fatigue reduction. This need was considered especially relevant as this fatigue was debilitating and interfered with patients’ daily functioning. Healthcare professionals reported that they currently have limited options to address severe fatigue, particularly after patients have already tried other forms of supportive care such as physiotherapy, occupational therapy or rehabilitation without sufficient improvement in fatigue symptoms.

*“[…] Patients often return unsatisfied after physio, occupational therapy, or rehab, in the sense that not all of their concerns have been resolved.” (Participant 023, Neurologist)*

With respect to the way in which the therapy is delivered, some psychologists questioned the added value of the fatigue-tailored content of bCBT, particularly in centers where psychologists already provide conventional CBT or Acceptance and Commitment Therapy to patients with glioma.

*“I did not get the impression that what I read about the fatigue-tailored bCBT was very different from what I already know and apply within cognitive behavioral therapy.” (Participant 013, Psychologist)*

#### Awareness as a prerequisite for adoption (knowledge and beliefs about the intervention, access to knowledge and information, structural characteristics, patient needs and resources)

### Symptom awareness

The understanding of fatigue varied across healthcare professionals. Many participants acknowledged the multifactorial nature of fatigue. However, some participants also found the underlying mechanisms of persistent severe fatigue difficult to understand, with several questioning whether fatigue was primarily related to neurological damage or tumor location.

*“I find it quite difficult to distinguish exactly why patients are struggling: whether it is purely fatigue, or also partly acquired brain injury. Maybe those are simply the same thing, I do not know.” (Participant 018, Nurse practitioner)*

An important determinant was whether fatigue was sufficiently recognized by healthcare professionals as a relevant symptom for identifying patients who may benefit from bCBT, especially as participants mentioned that patients do not always mention whether they are fatigued. Some of the nurse practitioners and neurologists indicated that fatigue was not routinely or explicitly assessed during consultations. Some neurologists asked about daily functioning frequently to assess patients’ activity levels, but did not specifically ask about fatigue while other neurologists tried to integrate it into their consultation but were not always able due to time constraints.

*“I do not explicitly ask about fatigue in every consultation, but I do ask what people do during the day, what they are able to do, and what they are not able to do. That often gives you an impression of their capacity.” (Participant 003, Neurologist)*

*“I notice that I still have to get used to incorporating PROMs [patient-reported outcome measures, red.] information into my follow-up consultations, where I only have 20 minutes and also need to discuss the MRI results and possible epilepsy.” (Participant 021, Neurologist)*

Regarding the other professionals, several psychologists indicated that fatigue was routinely assessed through the questionnaires used in their consultations. Nurse practitioners more often described fatigue as an important topic to address during clinical questioning, and when necessary relayed the information to the treating neurologists.

### Intervention awareness

Few participants were familiar with bCBT or had a clear understanding of the intervention. Awareness of the fatigue-tailored nature of bCBT was limited, particularly regarding its specific content and therapeutic components. Although many psychologists reported experience with CBT or other behavioral therapies, the specific fatigue-tailored elements of bCBT and how these differ from conventional CBT were generally unknown. Some neurologists and nurse practitioners indicated that they first learned about the content and mechanisms of the intervention during the interview. This limited awareness was identified as a barrier to implementation, as participants emphasized that even if bCBT were made available, it would be unlikely to be discussed with or offered to patients unless healthcare professionals were aware of the intervention and understood its relevance, especially with the many options that are already available.

*“I think everyone in the treatment team should be well informed about bCBT […] We should make sure that it is explained during tumor board meetings, so that everyone is aware of it and it becomes part of routine practice.” (Participant 021, Neurologist)*

#### Evidence as a lever for change (evidence strength and quality, knowledge and beliefs about the intervention)

Many healthcare professionals expressed a positive attitude toward bCBT, particularly because its effectiveness had been demonstrated in an RCT. Participants emphasized that bCBT represented one of the first evidence-based interventions specifically targeting severe fatigue in patients with glioma. This was often received positively, with participants describing a sense of relief that an intervention was available to address a symptom for which treatment options are currently limited.

*“Now there is something that has been proven in an RCT, one of the first truly evidence-based interventions that significantly improves fatigue in patients with diffuse glioma. In that sense, you can really make me excited about it. Now we finally have something.” (Participant 023, Neurologist)*

However, a few participants questioned whether evidence from a single RCT would be sufficient to support changes in clinical guidelines and some noted that colleagues might remain skeptical about the strength of the evidence.

*“However, one of my colleagues is very skeptical about bCBT. Of course, that is a valid perspective, but we view it quite differently.” (Participant 026, Nurse practitioner)*

#### Matching the intervention to the patient (relative advantage, complexity, patient needs and resources)

The complexity of identifying for which patients bCBT could be most suitable was frequently mentioned by neurologists and nurse practitioners. Participants described uncertainty about how bCBT should be positioned in relation to other available supportive care options, such as physiotherapy, occupational therapy or rehabilitation.

*“When you look at everything that is available online, there is so much that it is no longer clear what actually exists. For every complaint, it seems like patients can be referred somewhere.” (Participant 022, Nurse practitioner)*

Several participants reflected on whether bCBT would be appropriate for patients with high-grade gliomas, given the complexity of offering an intervention that requires time and effort in the context of a poor prognosis.

*“I can imagine that for patients with a higher-grade tumor, the consideration would be different. It is quite an investment, and the question is whether it will pay off in the time they still have. For patients with a low-grade tumor, that chance is obviously greater.” (Participant 016, Nurse practitioner)*

Despite these uncertainties, there was broad agreement that patients experiencing severe, persistent, and debilitating fatigue that interferes with daily functioning, would be suitable candidates for bCBT.

*“I think that when someone’s quality of life and sense of purpose are affected. […] I think bCBT should be introduced.” (Participant 017, Nurse practitioner)*

#### Pathways to psychological care (structural characteristics, implementation climate, compatibility)

Referral to psychological care varied across centers and was influenced by the existing organization of neuro-oncology services. In several centers, participants described close collaboration with psychological care within the hospital, particularly with neuropsychologists who were already involved in the assessment of patients with glioma. This preexisting connection was perceived as a potential facilitator for implementing bCBT, as some participants suggested that neuropsychologists may be well positioned to deliver or coordinate the intervention.

*“[…] But especially if the patient already knows them from an awake surgery, I think bCBT should be implemented there.” (Participant 023, Neurologist)*

However, the structure and availability of psychological care differed substantially between centers. While some neuro-oncology centers had dedicated neuropsychologists available for the assessment or treatment of patients with glioma, others reported limited or no options for referring patients to a psychologist within the hospital.

*“Availability is limited. Neuropsychology is a very large department, but it mainly focuses on diagnostics in general.” (Participant 003, Neurologist)*

Some psychologists also noted that, at present, few patients with glioma were referred specifically for severe fatigue. Current referrals were more often related to psychological concerns such as anxiety, depression, or adjustment difficulties. When fatigue was the primary complaint, participants indicated that patients were more commonly referred first to physiotherapy, occupational therapy or rehabilitation.

*“In practice, I see that patients are often referred to an occupational therapist or to rehabilitation for fatigue. They are usually not referred to me specifically for fatigue*.” *(Participant 012, Psychologist)*

#### Capacity constraints (available resources, structural characteristics, implementation climate)

Limited capacity within psychology departments was identified as an important barrier to implementation, particularly by psychologists. Several psychologists reported that existing services were already operating at full capacity, with waiting lists of several months in some centers. In this context, adding glioma patients to already constrained psychological care pathways was perceived as potentially challenging. However, some participants also noted that the expected number of eligible patients would likely be limited. If a small number of patients were to require bCBT, then it was generally considered manageable within existing capacity.

*“What I would wonder is: how many new patients would then come our way? We already have a waiting list of three to four months. If, for example, out of ten people referred to occupational therapy, one remains for whom it does not work sufficiently and who is then referred to the psychologist, then that may be feasible.” (Participant 012, Psychologist)*

Some neurologists and psychologists from certain centers mentioned that urgent cases, including patients with glioma, may be prioritized and therefore could receive care without following the standard waiting list. Some also mentioned that the waiting lists were not considered problematic as it was a matter of weeks instead of months before glioma patients could get treatment. The length of the waiting list and ability to prioritize glioma patients varied substantially across centers.

*“No, I do not think the waiting time would take very long. I think this could probably be arranged within two weeks. And if there were any waiting time, a patient with glioma would usually be prioritized if someone called about it.” (Participant 021, Neurologist)*

#### Funding the foundation (costs, available resources, compatibility)

Funding was identified as an important determinant for the implementation of bCBT, particularly by nurse practitioners and neurologists. Participants emphasized that implementation would be difficult if costs and financial responsibilities were not clearly defined from the outset. Moreover, several neurologists and psychologists questioned whether bCBT would be cost-efficient if it were used only for a relatively small subgroup of patients with glioma. For implementation to be considered worthwhile at the hospital level, some participants suggested that broader use across other cancer populations may be needed.

*“If the intervention is applicable not only to patients with brain tumors, but perhaps also to other patient groups, such as those with meningioma or brain metastases, then you would already have a somewhat larger group.” (Participant 003, Neurologist)*

Psychologists perceived costs for bCBT as manageable, noting that funding could be available for the required training and that the remaining time investment would fall within psychologists’ regular working hours.

*“As a neuropsychology department, we are fully embedded within the cost pathway of the neurologists and neurosurgeons. Honestly, many of our activities are already included within that structure, so it would be very unlikely for this to generate additional costs.” (Participant 024, Psychologist)*

However, the digital platform subscription required for bCBT was identified as a specific barrier by several psychologists. Many hospitals used a digital platform different from the one used in the RCT, and participants questioned whether hospitals would be willing to finance an additional platform subscription solely for bCBT.

### Proposed implementation strategies

Implementation strategies were developed to overcome the identified barriers. All strategies, strategy type, barriers addressed and responsible healthcare professional are shown in Table 3. The strategies are presented in the three phases representing the clinical pathway. The phases are explained below and correspond with the implementation roadmap shown in Figure 1, which was developed as a supportive tool for implementation.

**Figure 1.**
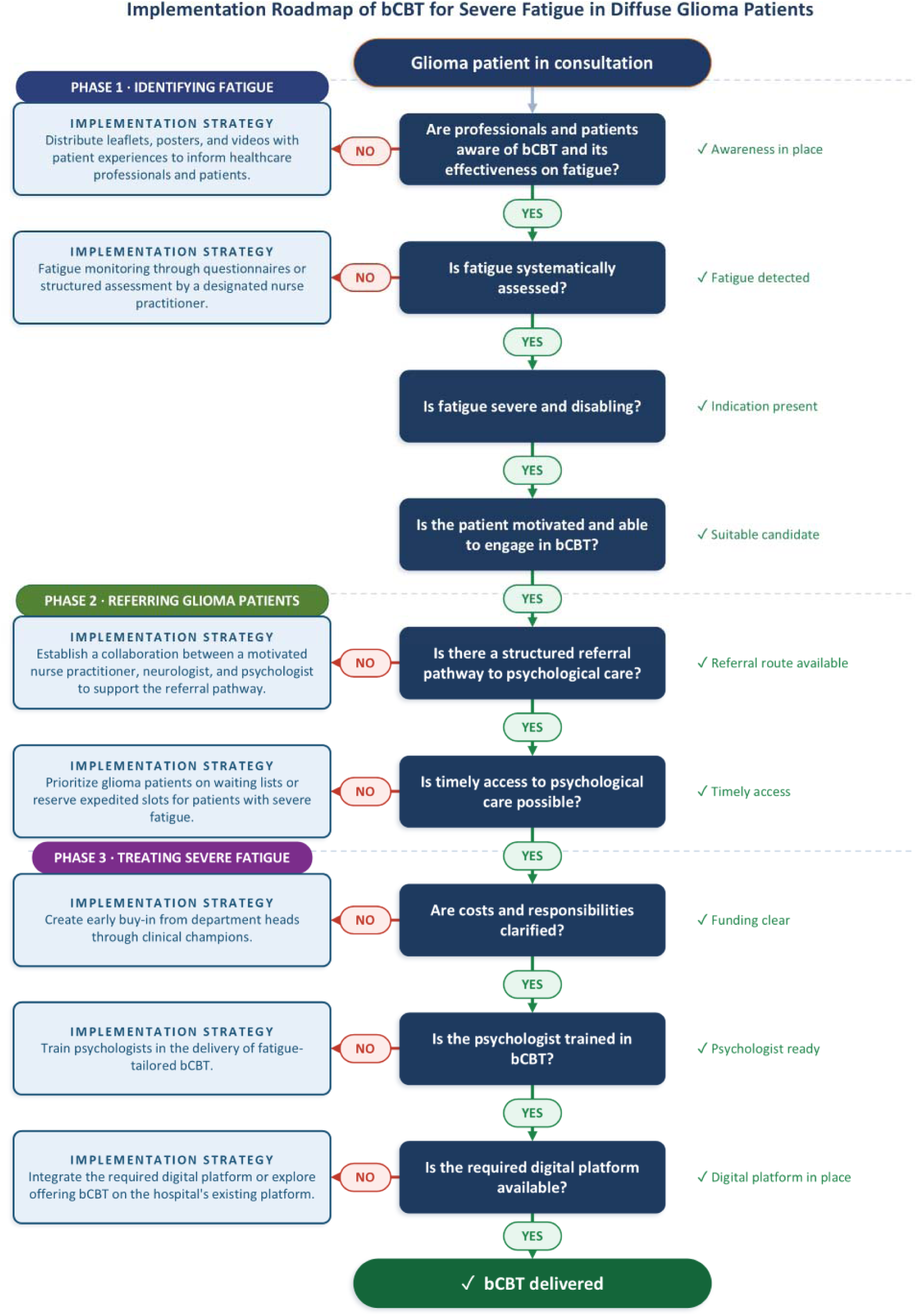
Implementation roadmap representing the three phases of the clinical care pathway for glioma patients: Phase 1: Identifying fatigue, Phase 2: Referring glioma patients and Phase 3: Treating severe fatigue. The implementation roadmap is a supportive tool with strategies for every phase of the clinical pathway. Abbreviations: bCBT = blended cognitive behavioral therapy.

**Table 3.** Implementation strategies organized by the barriers addressed, the responsible healthcare professional, and structured by phase of the clinical pathway.

|  | Strategy type | Implementation strategy | Barriers addressed | Responsible healthcare professional |
|---|---|---|---|---|
| Phase 1: Identifying fatigue | Informational | Promotion and distribution of leaflets, posters, and videos with patient experiences | • Limited familiarity with bCBT and its fatigue-tailored content<br>• Underlying mechanisms of fatigue considered difficult to understand<br>• Unclear added value over existing CBT<br>• Uncertainty about positioning bCBT relative to other options<br>• Anticipated skepticism among colleagues<br>• Patients do not always mention fatigue | Nurse practitioners, neurologists, psychologists |
|  | Organizational | Fatigue monitoring | • Fatigue not routinely assessed<br>• Patients do not always mention fatigue | Nurse practitioners |
| Phase 2: Referring diffuse glioma patients | Collaboration-promoting | Establishing collaboration between a motivated nurse practitioner, neurologist, and psychologist to support the referral pathway | • Limited in-hospital referral option<br>• Fatigue rarely the primary referral reason | Nurse practitioners, neurologists, psychologists |
|  | Organizational | Prioritize glioma patients or reserve expedited slots on waiting lists | • Psychology departments at full capacity<br>• Waiting lists of several months | Psychologists |
| Phase 3: Treating severe fatigue with bCBT | Motivational | Create early buy-in from department heads through clinical champions | • Implementation difficult without clear cost arrangements<br>• Cost-efficiency questioned for a small subgroup | Clinical champions |
|  | Educational | Train psychologists in the delivery of fatigue-tailored bCBT | • Limited familiarity with bCBT and its fatigue-tailored content<br>• Unclear added value over existing CBT<br>• Underlying mechanisms of fatigue considered difficult to understand | Psychologists |
|  | Facilitative | Integrate the required digital platform | • App subscription used (Therapieland vs. MindDistrict) | Clinical champions |

Note. Abbreviations: bCBT = blended cognitive behavioral therapy; CBT = cognitive behavioral therapy.

### Phase 1: Identifying fatigue

*Promotion and distribution of leaflets, posters, and videos with patient experiences.* This strategy is proposed to spread awareness of bCBT and clarify how bCBT differs from other commonly used interventions for severe fatigue. Promotional materials could also encourage patients to discuss fatigue with their healthcare professionals.

Fatigue monitoring. Making a designated healthcare professional, such as a nurse practitioner, responsible for systematically assessing fatigue could improve the identification of severely fatigued patients and allow referral to bCBT to be considered. Fatigue can either be assessed through questionnaires if feasible within clinical workflows or through structured clinical assessment.

### Phase 2: Referring diffuse glioma patients

*Establishing collaboration between a motivated nurse practitioner, neurologist, and psychologist to support the referral pathway.* Collaboration between the three disciplines could support the referral procedures of bCBT and improve access to broader psychological care.

*Prioritize glioma patients or reserve expedited slots on waiting lists.* Where waiting lists extend to several months, prioritizing glioma patients or reserving expedited slots could allow timely access to psychological care.

### Phase 3: Treating severe fatigue with bCBT

*Create early buy-in from department heads through clinical champions.* Clinical champions, healthcare professionals who support bCBT and are willing to lead its implementation, could engage department heads. This can help clarify financial and organizational responsibilities for training costs, therapist time and platform access.

*Train psychologists in the delivery of fatigue-tailored bCBT.* Training may address the uncertainty about the relative advantage of the fatigue-tailored content while also increasing psychologists’ knowledge of the intervention and its appropriate use in glioma patients.

*Integrate the required digital platform.* Centers could integrate the bCBT platform or bCBT could be made available on platforms already in use.

## Discussion

This study aimed to identify barriers and facilitators to implementing bCBT for severe fatigue among patients with glioma in routine hospital care. Guided by the CFIR, key identified themes included the perceived need for fatigue reduction with bCBT, time and financial constraints, awareness of fatigue and bCBT, patient suitability for bCBT and confidence in the intervention’s evidence base and effectiveness. To address these, seven implementation strategies were proposed across three phases of the clinical pathway.

Previous implementation research on CBT and internet-based CBT (iCBT) has similarly identified resource constraints and organizational factors influencing uptake in clinical practice.^25,35–38^ The present study extends these findings to the neuro-oncology context. Our findings are also consistent with previous research on cancer-related fatigue in the Netherlands, which showed that even guideline-recommended care does not consistently reach patients.^37^ This was partly due to limited knowledge of interventions or their suitability and unclear responsibility for identifying and managing fatigue.^37^ In our study, nurse practitioners more often addressed fatigue during consultations and relayed this information to the treating neurologist, and they therefore may be best positioned to take ownership of fatigue identification and referral. Furthermore, clinical champions may also be important to support awareness, and integration of bCBT into routine hospital care by engaging department heads.^33,34,39^ The most suitable champion may depend on which department carries the financial responsibility.

Across participants, fatigue was generally understood as a multifactorial symptom. However, fatigue was also frequently attributed to acquired brain injury, which may lead to underrecognition of the cognitive, behavioral, and emotional factors that can contribute to persistent severe fatigue in some patients with glioma.^14^ From a clinical perspective, this may result in severely fatigued patients being primarily referred to supportive care options such as physiotherapy, occupational therapy, or rehabilitation, even though these patients may not sufficiently benefit from such referrals.^40^ Therefore, bCBT should be positioned as a targeted treatment option for severe persistent fatigue in glioma alongside the existing referral options, rather than as an intervention only considered after all other options have failed. Patients whose fatigue is maintained by factors addressed within the bCBT protocol, including maladaptive cognitions, anxiety-related concerns, disrupted sleep, and unbalanced activity patterns, may be particularly suitable.^14^ In these cases, referral to bCBT may be more appropriate than default referral to occupational therapy or rehabilitation.^14,40^ Nevertheless, there is no literature on when which treatment would be most beneficial for fatigue in glioma patients. Further research should inform which evidence-based treatment to use and when.

Similarly, from a psychological care perspective, there may be a tendency to prioritize anxiety or depression when these symptoms are present, as indicated by several psychologists in our study. However, because fatigue can also be understood as partly maintained by psychological and behavioral factors, it may be appropriate in some cases to address severe fatigue first rather than automatically prioritizing anxiety or depressive symptoms.^13,14^ This would require a shift in focus from current referral and treatment routines. However, this approach should not be applied indiscriminately. Treatment priorities should be determined on an individual basis, taking into account the patient’s severity of other psychological symptoms, cognitive functioning and preferences. It is also important to acknowledge that a proportion of patients may remain severely fatigued despite bCBT.^13^ For these patients, additional or alternative approaches may be needed.

Beyond the proposed strategies, several considerations remain relevant for implementation. Firstly, when questionnaires such as the Checklist Individual Strength or other Patient-Reported Outcome Measures (PROMs) are used for fatigue monitoring, results should be discussed with patients.^41,42^ Previous literature and clinical experience suggest that patients may become less motivated to complete questionnaires when responses are not addressed or feedback is absent.^43–46^ Routine monitoring with PROMs may also offer broader benefits as it has been associated with improved HRQoL and overall survival in other oncological populations.^47,48^ Secondly, timely access to psychological care is particularly relevant, as waiting times of several months may represent a substantial proportion of the remaining life expectancy of some patients.^18^ Furthermore, suitability for bCBT in patients with high-grade glioma should be assessed individually, as their relatively poor prognosis makes referral complex.^18,19^ However, fatigue reduction and regaining a sense of control may remain meaningful goals for some patients, including in the context of limited life expectancy.^6,7,18,24^ Therefore, bCBT should not be excluded solely based on tumor grade and may be discussed when fatigue reduction is a patient priority and the intervention is feasible. Lastly, financial feasibility may be improved if fatigue-tailored bCBT is implemented more broadly. Evidence suggests that CBT targeting severe fatigue may have transdiagnostic potential and transdiagnostic training is already available.^20–24,49^ However, further research is needed to determine whether shared training is sufficient or whether disease-specific adaptations and supervision remain necessary.

This study had several strengths and limitations. A limitation is that patients were not interviewed. As a result, patient needs and preferences were explored indirectly through the perspectives of healthcare professionals. This may have limited insight into when patients themselves would raise fatigue as a problem, how they would perceive bCBT, and whether they would be willing to use the intervention. However, previous research has shown that at least a subgroup of patients with glioma is willing to engage in psychoeducation for their fatigue.^6,7^ This was also reflected in the patient evaluation of the bCBT trial in diffuse glioma patients. Overall acceptance was high and the majority considered the intervention useful and supportive despite the intensity of the program.^13^ Moreover, the primary aim of this study was to identify barriers and facilitators within the hospital implementation context. Healthcare professionals were therefore considered the most suitable participants. With the inclusion of 26 healthcare professionals, a rich dataset was provided encompassing multidisciplinary perspectives of professionals involved in the care pathway relevant to bCBT implementation. However, future work should also include the perspectives of oncologists, radiotherapists, social workers and patients as this could provide additional information.

A strength of this study is that it combined a pre-implementation assessment with the development of implementation strategies tailored to the barriers identified in the interviews. Although previous studies have examined barriers and facilitators to the implementation of CBT and iCBT, fewer studies explicitly translate these findings into context-specific strategies and none within the pre- implementation stage.^35,36,38^ This added a practical dimension to the study by moving beyond problem identification toward implementation planning. Although this context-specific approach may have limited the direct transferability to healthcare systems in other countries, it increased the practical relevance of the proposed strategies for Dutch neuro-oncology care. Future implementation research could consider combining ERIC-based strategy selection with context- specific frameworks and stakeholder input when aiming for broader national implementation.^33^ In addition, the implementation roadmap presented in Figure 1 provides a practical supporting tool for centers.

The road to implementation may differ between centers. In some centers, implementation of bCBT might be feasible as only a limited number of strategies may be needed, as several components of the clinical care pathway needed to support bCBT implementation are already in place. However, in other centers several strategies may need to be applied simultaneously. This could complicate implementation, especially if bCBT is to be introduced within a short timeframe. In these cases, an implementation pilot may be beneficial. One approach could involve a collaboration between a center in which bCBT for fatigue is already implemented, and one or more participating centers. This would allow potential barriers to be addressed in a structured way while maintaining intervention quality and ensuring that appropriate patients are included. If the pilot is successful, broader and independent adoption by the participating center should be considered as a next step. Additional long-term implementation options could then be explored, such as training rehabilitation psychologists rather than medical or neuropsychologists, particularly in centers where capacity constraints within psychological care are a major barrier.

To conclude, this study identified multiple themes relevant to implementing bCBT for severe fatigue in patients with diffuse glioma. Implementation will likely require center-specific strategies that align with existing workflows and organizational structures. A phased approach focusing on fatigue identification, referral of suitable patients, and treatment delivery may support the integration of bCBT into routine neuro-oncology care.

## Supplementary data

Supplementary Figure 1 consists of the topic list for healthcare professionals made with the CFIR framework. Supplementary Figure 1 was added as a PDF file with the submission of this document. The COREQ Checklist is added as a PDF file with the submission of this document.

## Author Contributions

Conceptualization and design: I.E., M.G., L.D., M.K., and M.S. Participant recruitment: I.E., M.G., J.M.N., and M.K. Data collection and curation: I.E., and M.G. Data analysis and interpretation: I.E., M.G., L.D., M.K., and M.S. Visualization: I.E. Writing of manuscript: I.E. and M.G. Revising of manuscript: all authors. Final approval of manuscript: all authors. Accountable for all aspects of the work: all authors.

## Supporting information

COREQ checklist

## Data Availability

All data produced in the present study are available upon reasonable request to the authors

## Ethics Statement

This study was approved by the ethical review board of the Amsterdam University Medical Center (METc 2026.0223). All participants provided written informed consent prior to study participation.

## Data Availability

Data will be made available upon reasonable request.

