## Supplementary Figure 1 for "Implementing blended cognitive behavioral therapy for severe fatigue in routine hospital care for diffuse glioma patients: a qualitative study"

### Topic list for healthcare professionals

Interviewer(s):
Date:
File number:

Introduction:

Thank you for participating in this study. My/our name(s) is/are […]. This study is set up by our GRIP team, which focuses specifically on fatigue experienced by glioma patients. The main topic of today is about implementing blended cognitive behavioral therapy to reduce severe fatigue in diffuse glioma patients in the standard hospital care. bCBT consists of conventional CBT and online modules with therapist feedback. It is a 12-week program with 5 patient-therapist sessions either face-to-face or via video consult. This variant enhances accessibility, flexibility and engagement while improving treatment efficacy.

The purpose of this interview is to gain insight on your views on bCBT for this patient population and in your medical center. The goal of the study is to identify the barriers and facilitators of the intervention to optimize the implementation process in multiple centers.

I/we would like to ask your permission to record the interview for transcription and evaluation afterwards by the team. The interview will last approximately 60 minutes. The interview will remain confidential and will only be used for this study.

Do you have any questions before we begin?

| **Demographic Characteristics Questions** | **Answers** |
|---|---|
| What is your age? | … years |
| What is your gender? | Man / Woman/ Other |
| At which (healthcare) institution are you employed? | |
| What is your position? | |
| How many years have you worked in this position? | … year(s) |

| Domain: Intervention | Question |
|---|---|
| Construct: relative advantage | What do you think of bCBT as a treatment for fatigue in glioma patients?<br><br>Prompt: How do you think bCBT compares to the standard of care for fatigue in glioma patients?<br><br>Prompt: Do you think there are better alternatives and if so, in what way are they better? |
| Evidence strength and quality | What do you know about bCBT?<br><br>Prompt: What is your opinion on bCBT or behavioral treatments in general? |
| Complexity | What do you think of bCBT as an intervention?<br><br>Prompt: How complicated do you think the intervention will be for your colleagues and patients? |
| Cost | Do you have any idea what the costs will be for the implementation and usage of bCBT? |
| **Domain: Outer setting** | |
| Patient needs and resources | Do patients mention fatigue to you?<br><br>Prompt: Do you ask about fatigue or do the patients mention it themselves?<br><br>Prompt: Do you think there is a need to reduce fatigue in this patient population? |
| | Are there any factors such as costs, rooms or time, that would limit or facilitate the usage of bCBT by you and your colleges? |

| | |
|---|---|
| | What is in your opinion the best strategy to reduce fatigue in glioma patients?<br><br>Prompt: Do you think the bCBT will meet the need of fatigue reduction in patients? |
| | How do you think patients will interact with or respond to bCBT?<br><br>As you might know, the patients need to do online modules at home and interact with the therapist feedback.<br><br>Prompt: How do you see this patient population engaging in the online modules of the intervention? |
| | How would patient costs be reimbursed or covered? |
| **Domain: Inner setting** | |
| Structural characteristics | How is psychological care organized in your medical center?<br><br>Prompt: Are there any limitations, such as referral systems or understaffing, within the organization of psychological care for the use of bCBT?<br><br>Prompt: If so, what is needed to solve this within your organization? |
| Implementation climate<br>Subconstruct: Compatibility | What do you do in the case of fatigue symptoms in patients?<br><br>Prompt: Who do you think should deliver bCBT?<br><br>Prompt: How well does the intervention fit in the current work processes/practices and referral system? |

| | |
|---|---|
| | How important do your colleagues consider fatigue as a symptom in diffuse glioma patients. |
| Readiness for implementation | A key factor for successful implementation is the setting's commitment to and support of the intervention.<br><br>What do your colleagues think of bCBT and would they propose/deliver bCBT to glioma patients?<br><br>Prompt: If so, what would facilitate this support? If not, why would they not support bCBT? |
| Available resources | What programs and treatments are already present similar to bCBT for this patient population?<br><br>Prompt: Would the presence of these programs and treatments hinder or aid the implementation of bCBT in any way? |
| | Approximately how many patients with severe fatigue symptoms do you encounter in your practice?<br><br>Prompt: How many staff members do you think would be required to deliver this intervention effectively to a patient?<br><br>Prompt: Is there enough staff for the usage of bCBT for all glioma patients with severe fatigue in your setting right now? |
| | Are there factors, such as costs, space, or time, that would limit or facilitate the use of bCBT by you and your colleagues? |
| Access to knowledge and information | The intervention consists of fatigue tailored-blended cognitive behavioral therapy that is delivered by psychologists. |

| | |
|---|---|
| | How can we ensure that the psychologists at your center receive appropriate training?<br><br>Prompt: How is knowledge kept up to date within your center, and would you and your colleagues be open to attending a training at Amsterdam UMC? |
| **Domain: Characteristics of individuals** | |
| Knowledge and beliefs about the intervention | Do you have any affinity with psychotherapy?<br><br>Prompt: Why not?<br>Why so? |
| | Would you propose/deliver bCBT to a patient presenting with severe fatigue symptoms?<br><br>Prompt: If not, why would you not?<br>If so, why? |
| | What do you think about the symptom fatigue in glioma patients?<br><br>Do you consider fatigue a consequence of treatment/disease?<br><br>Prompt: What do you think about the persistence of fatigue in the patient population? |
| | Do you think bCBT will be effective in your setting?<br><br>Prompt: Why not?<br>Why so? |
| Self-efficacy | How confident are you that you will be able to give bCBT to glioma patients? |

| Domain: Implementation process | |
|---|---|
| Planning | What do you think is important to consider during the implementation in your center?<br><br>Prompt: And how about nationally? |
| | Do you expect that the intervention will be easy to implement in your setting?<br><br>Prompt: If so, what would facilitate the implementation?<br>If not, what would be the major obstacles? |
| Engaging<br>Subconstructs: external change agents | Is there a need for an outside organization to help with the implementation? |
| Subconstruct: Intervention participants | If bCBT would be available, how would you propose the intervention to patients?<br><br>Prompt: How would you describe the intervention to patients?<br><br>Prompt: How would you monitor fatigue and when would you propose this option? |
| | How will patients access the intervention?<br><br>Prompt: Would patients be able to access bCBT immediately?<br><br>Prompt: Would there be a waiting list for the patients?<br><br>Prompt: If so, how long? |
| **Finalization of interview** | |
| | Summarize the results of the interview to the interviewee. |

| | |
|---|---|
| | Is there anything that you would like to add or comment about which has not been discussed yet? |
